# Mortality among patients with invasive group A streptococcal infections caused by the M1_UK_ lineage: a retrospective cohort study in England and Wales

**DOI:** 10.1101/2025.06.25.25330084

**Authors:** Ho Kwong Li, Nina Zhu, Olivia Waddell, Juliana Coelho, Roger Daniel, Rebecca L Guy, Theresa Lamagni, Shiranee Sriskandan

## Abstract

**Background:** The M1_UK_ sublineage of *S. pyogenes* has dominated post-pandemic upsurges in invasive group A streptococcal (iGAS) disease. Using a national cohort of patients with *emm*1 iGAS infections, we compared the case fatality rate (CFR) attributable to M1_UK_ with ancestral *emm*1 lineages.

**Methods:** We linked *emm*1 *S. pyogenes* iGAS cases in England and Wales between December 2009-July 2022 to demographic and mortality data. We assigned lineage to isolates from 2010, 2013-2016, and 2020 using genome-sequencing or allele-specific PCR. Seven and 30-day all-cause CFR were determined for the entire cohort. Univariate and multivariate analyses were conducted to determine if lineage affected risk of death.

**Results:** In total, 4,952 *emm*1 iGAS cases were linked with demographic and mortality data; lineage was assigned to 1,356 cases. M1_UK_ was associated with a 30-day CFR of 24.4%, compared with 22.3% for M1_global_, 10.5% for M1_23SNP_, and 10.3% for M1_13SNP_. After adjustment for age and sex, *emm*1 lineage was not a significant risk factor for death within 7 or 30 days. Survival analysis highlighted rapid time to death as a feature of both M1_UK_ and M1_global._ Most deaths (63.7%) occurred within 1d of diagnostic sampling. Of children under 15 who died, 56.3% died before any sampling, and 95.6% died within 1d of sampling.

**Conclusions:** Significant differences in mortality were not identified between M1_UK_ and M1_global_ but larger studies are required. Mortality due to *emm*1 *S. pyogenes* is exceptionally high. Rapid time to death has implications for clinical trials: to impact death, interventions are required prior to culture-based diagnosis.

## Introduction

Though rare, invasive group A Streptococcal (iGAS) infection can be associated with significant mortality and devastating sequelae for those affected. Manifestations include bacteraemia, pneumonia, skin and soft tissue infection including necrotizing fasciitis, puerperal sepsis and streptococcal toxic shock (1, 2). Since relaxation of non-pharmaceutical interventions (NPI) to combat the spread of SARS-CoV2, upsurges in iGAS infections have been reported by national public health institutes and networks in Europe, North and South America, Australia, New Zealand, and Japan, some reporting high mortality (3–14). Waning population immunity and consequent rebound upsurges of seasonal respiratory viruses were predicted following cessation of COVID-19-related NPI, although bacterial infections such as *Streptococcus pyogenes* were notably omitted (15). Many, though not all, *S. pyogenes* upsurges have reported a predominance of an emergent lineage of genotype *emm*1, M1_UK_. (6–9,14,16–17) The M1_UK_ *S. pyogenes* lineage is distinguished from ancestral *emm*1 strains by 27 core SNPs and constitutive production of the erythrogenic streptococcal pyrogenic exotoxin A (SpeA) (18). Emerging around 2008, M1_UK_ has almost replaced ancestral strains of *emm*1 in the United Kingdom representing 97% of *emm*1 (16), and currently represents a majority of *emm*1 isolates circulating in several European countries, Japan, Canada, New Zealand, and Australia (6–9, 11, 14,16–20).

Post-pandemic upsurges in iGAS have differed between countries, potentially linked to different timing of NPI ending, population structure, season, and climate. Several European countries reported an initial increase in *emm*12 infections followed by *emm*1 (5,8). A notable increase in frequency of paediatric empyema was reported in Europe, over 80% of which were linked to M1_UK_ infections in the United Kingdom (22). Mortality among children in England with iGAS lower respiratory tract infections in the period October-December 2022 was very high (25%), associated with respiratory viral co-infection (23). Despite different seasons and prevailing viruses, an increase in paediatric iGAS infections was also observed in Australia (12). In contrast, in Japan, where NPI were relaxed later than the United Kingdom, cases of streptococcal toxic shock were reported to increase in 2023-24, predominantly affecting adults, although also associated with a predominance of the M1_UK_ lineage (14).

Overall mortality from iGAS in developed countries like the USA is reported to be 9.3% (1), but mortality is known to be influenced by individual host susceptibilities such as age, and by causative strain genotype (2, 24). *emm*1 and *emm*3 infections are acknowledged to be associated with increased risk of severe outcomes and mortality compared with other *emm* genotypes, although most large-scale population-based epidemiological studies were undertaken prior to the emergence of M1_UK_ (2, 24). Invasiveness and virulence of *S. pyogenes* has been attributed in part to a propensity to undergo mutations in the two component regulator covRS during the transition from non-invasive to invasive infection, resulting in de-repression of key virulence genes (25). Although most experimental work relating to covRS has focused on *emm*1 (25), all *emm* genotypes are capable of similar mutation during clinical invasive infection (26). Hence, it may be that other constitutive features unique to the modern clone of *emm*1, including M1 protein itself, the streptococcal inhibitor of complement (SIC), phage-encoded toxins and DNAses, increased production of streptolysin O, are additionally important (27–28).

Early reports of M1_UK_ association with increased iGAS frequency (18) and case severity (29), prompted us to determine if the mortality associated with M1_UK_ iGAS differed from that associated with ancestral *emm*1 isolates. Ancestral isolates included the previously dominant ‘pandemic’ M1_global_ strains, and two intermediate sublineages M1_13SNP_ and M1_23SNP_, which preceded the emergence of M1_UK_ (16). The study included all *emm*1 iGAS cases in England and Wales where *emm*1 lineage had been assigned by genome sequencing or allele specific PCR. To include sufficient iGAS cases associated with M1_global_ infection, we extended genotyping to include cases from 2010, while, to avoid any potential effect of population-level changes in anti-streptococcal immunity on case fatality, cases after 2020 were not included in lineage-based analyses.

## Methods

### Demographic data for patients with *emm*1 iGAS 2009-2022

Demographic data (age, sex, ethnicity, index of deprivation) and mortality of all patients who acquired invasive *emm*1 *S. pyogenes* infection between December 2009 and July 2022 in England and Wales were obtained by linking national bacteriology data held by UKHSA to individual NHS records. Sterile site *S. pyogenes* isolates are sent from all laboratories in England and Wales to the UKHSA *Staphylococcus* and *Streptococcus* Reference Section (SSRS); these isolates are characterized using *emm* gene sequencing. Patient records from Dec 2009-July 2022 with *emm*1.0 isolates were submitted to the NHS Demographic Batch Tracing Service to identify date of death (30). For unsuccessfully traced cases a further assessment against the NHS Personal Demographics Service (PDS) was used to identify the date of death (31). For mortality analysis, patients were considered lost to follow up if they could not be successfully traced to the NHS SPINE through either method. Demographic and ethnicity data were obtained through linkage to Hospital Episode Statistics (HES) using unique NHS number and date of birth (32). Ethnicity was allocated following the COVID-19 Health Inequalities Monitoring for England (CHIME) tool methodology (33). UK Health Security Agency surveillance of infections for health protection purposes is approved under Regulation 3 of The Health Service (Control of Patient Information) Regulations 2020 and under Section 251 of the NHS Act 2006.

### Emm1 lineage assignment

Lineage assignment was undertaken for all *emm*1 iGAS isolates previously sequenced between 2013 to 2016 and for all iGAS *emm*1 from 2020 analysed using allele-specific polymerase chain reaction (AS-PCR) (18, 34). Lineage was assigned for all *emm*1 iGAS isolates from the year 2010 using AS-PCR (34) as part of this work.

### Statistical analysis

We defined 30-day all-cause mortality to include a documented death date falling between 7 days before and 30 days after the specimen date (or 7 days before and 7 days after specimen date for 7-day mortality) and calculated the case fatality rate accordingly, to accommodate the fact that some diagnostic samples were obtained postmortem. Univariate and multivariate analysis to determine the impact of lineage on outcome was limited to those patients with *emm*1 iGAS for whom lineage was determined and where linkage to NHS data sources was successful.

Univariate analysis was performed to analyse variation in odds ratio of mortality in different sub-groups using the X^2^ or Fisher exact test for categorical variables, and Student t-test for continuous variables. The independent variables tested for univariate association with death included age, sex, ethnicity, socioeconomic deprivation, and sample year. Sample proportions, *i.e.*, the case-fatality rate (CFR), defined as number of deaths in iGAS cases of interest divided by the total number of iGAS cases in the same group of interest (for example M1_UK_ or M1_global_), were compared using z-test. A multivariate model of predictors of 7-day and 30-day all-cause mortality for cases with lineage assigned was developed. All reported statistical tests were 2-sided, and p-values <0.05 were considered statistically significant. We performed survival analysis using specimen date as day 0, and a half-day interval to better differentiate those who died before culture confirmation (i.e. specimen obtained post-mortem) from those who died on the same day as specimen date. As such, patients with a sample (that confirmed *S. pyogenes*) taken up to 7 days after death were assumed to have survived 0 days, while patients who died on the day that the sample was taken were assumed to have survived 0.5 days. Kaplan-Meier survival analysis was undertaken for the first 30 days after an *emm*1 isolate was identified, comparing age groups, and *emm*1 lineages (35) and compared using the log rank test (36,37).

## Results

### Study cohort, isolates, and *emm*1 lineages

Between December 2009 and July 2022, 5,274 *S. pyogenes emm*1 isolates were identified from the UKHSA’s SGSS national surveillance hub. (Supplementary figure S1A).

After excluding 254 isolates that were not from England and Wales, or were duplicates, 5,020 *emm*1 isolates were linked with patient ethnic group data in HES and survival/death information in DBS. Linkage was successful for 99% (n = 4,952) of these *emm*1 cases.

Overall, 1,445 *emm1* isolates were assigned a lineage, however 67 of these were excluded as they were not from England and Wales. Of the 1,378 invasive *emm*1 isolates assigned to a lineage, 98.4% (n = 1356) were linked to demographic and mortality data, accounting for 27.8% (1356 / 4952) of all *emm1* invasive isolates. These *emm*1 isolates comprised 39.1% M1_global_ strains (n = 530); 57.4% M1_UK_ strains (n = 778); 2.1% M1_13SNP_ strains (n = 29); and 1.4% M1_23SNP_ strains (n = 19). As expected, M1_UK_ strains became more frequent over time, accounting for 0.27% (1/375) eligible cases with lineage-assigned in 2010, and 90.1% in 2020 (201/223) (Supplementary figure S1B). Three-quarters (203/268) of eligible *emm*1 isolates from 2020 were from the first three months of the year, prior to the main impact of the COVID-19 pandemic. (Supplementary Figure S1a).

### Case fatality rate

Overall, 1,043 deaths were recorded within 30 days among the 4,952 *emm*1-infected iGAS patients, yielding a 30-day case fatality rate (CFR) of 21.1% (95% CI 19.9%, 22.2%).

The 7-day CFR was 18.2% (95% CI: 17.1%, 19.3%) (Table 1). The 30-day CFR rose markedly with age, rising from 13.1% in children under 15 years, to 49.5% in the over 85 years group (Supplementary Table S1). The CFR did not change when excluding the period of COVID-19-related lockdowns (23 March 2020 to 23 February 2021) 30-day CFR: 21.0% (95% CI: 19.8%, 22.1%); 7-day CFR: 18.1% (95% CI: 17.0%, 19.2%), however, the numbers of *emm*1 iGAS during this period were very limited.

**Table 1.** Patient characteristics and univariate analysis.

|  |  | Number (N) | Percentage (%) | Mortality, 30 days, odds ratio (95% CI) | p-value | Mortality, 7 days, odds ratio (95% CI) | p-value |
| --- | --- | --- | --- | --- | --- | --- | --- |
| Total |  | 4,952 |  |  |  |  |  |
| All age |  |  |  |  |  |  |  |
| Gender | Male | 2,608 | 52.67 | Reference |  | Reference |  |
|  | Female | 2,317 | 46.79 | 1.16 (1.01, 1.33) | 0.04 | 1.16 (1.01, 1.34) | 0.04 |
|  | Unknown | 27 | 0.55 |  |  |  |  |
| Age group | Under 15 | 950 | 19.18 | Reference |  | Reference |  |
|  | 16 to 64 | 1,887 | 38.11 | 1.15 (0.91, 1.44) | 0.24 | 1.01 (0.8, 1.27) | 0.95 |
|  | 65 to 84 | 1,606 | 32.43 | 2.14 (1.72, 2.67) | 0.00 | 1.66 (1.32, 2.08) | 0.00 |
|  | 85 and above | 505 | 10.20 | 6.53 (5.05, 8.44) | 0.00 | 4.67 (3.6, 6.04) | 0.00 |
|  | Unknown | 4 | 0.08 |  |  |  |  |
| Age (year) |  |  |  | 1.02 (1.02, 1.02) | 0.00 | 1.02 (1.01, 1.02) | 0.00 |
| Ethnicity | White | 3,895 | 78.66 | Reference |  | Reference |  |
|  | Asian | 296 | 5.98 | 0.77 (0.56, 1.05) | 0.10 | 0.88 (0.63, 1.21) | 0.42 |
|  | Black | 116 | 2.34 | 0.74 (0.45, 1.22) | 0.24 | 0.85 (0.51, 1.42) | 0.54 |
|  | Mixed | 59 | 1.19 | 0.87 (0.45, 1.68) | 0.68 | 1.06 (0.55, 2.06) | 0.86 |
|  | Other | 33 | 0.67 | 1.02 (0.44, 2.36) | 0.96 | 1.25 (0.54, 2.89) | 0.60 |
|  | Unknown | 553 | 11.17 |  |  |  |  |
| IMD | 1 | 746 | 15.06 | Reference |  | Reference |  |
|  | 2 | 638 | 12.99 | 0.86 (0.66, 1.11) | 0.25 | 0.93 (0.71, 1.23) | 0.63 |
|  | 3 | 692 | 13.97 | 0.89 (0.69, 1.14) | 0.35 | 0.92 (0.70, 1.21) | 0.56 |
|  | 4 | 680 | 13.73 | 0.94 (0.73, 1.21) | 0.62 | 1.10 (0.84, 1.43) | 0.48 |
|  | 5 | 618 | 12.48 | 1.08 (0.84, 1.39) | 0.56 | 1.07 (0.81, 1.41) | 0.63 |
|  | Unknown | 1,578 | 31.87 |  |  |  |  |
| Lineage | M1<sub>global</sub> | 530 | 10.70 | Reference |  | Reference |  |
|  | M1<sub>13</sub>SNP | 29 | 0.59 | 0.40 (0.12, 1.35) | 0.14 | 0.47 (0.14, 1.59) | 0.23 |
|  | M1<sub>23</sub>SNP | 19 | 0.38 | 0.41 (0.09, 1.8) | 0.24 | 0.23 (0.03, 1.72) | 0.15 |
|  | M1<sub>UK</sub> | 778 | 15.71 | 1.13 (0.87, 1.47) | 0.37 | 1.12 (0.85, 1.47) | 0.42 |
|  | Unknown | 3,596 | 72.62 |  |  |  |  |
| Sample year | 2009 | 4 | 0.08 | Reference |  | Reference |  |
|  | 2010 | 396 | 8 | 0.92 (0.09, 8.96) | 0.94 | 0.77 (0.08, 7.51) | 0.82 |
|  | 2011 | 374 | 7.55 | 0.82 (0.08, 7.95) | 0.86 | 0.67 (0.07, 6.51) | 0.73 |
|  | 2012 | 387 | 7.82 | 0.98 (0.10, 9.49) | 0.98 | 0.86 (0.09, 8.35) | 0.89 |
|  | 2013 | 430 | 8.68 | 0.82 (0.08, 7.94) | 0.86 | 0.65 (0.07, 6.38) | 0.72 |
|  | 2014 | 352 | 7.11 | 0.83 (0.08, 8.06) | 0.87 | 0.73 (0.07, 7.14) | 0.79 |
|  | 2015 | 494 | 9.98 | 0.91 (0.09, 8.84) | 0.94 | 0.76 (0.08, 7.4) | 0.81 |
|  | 2016 | 590 | 11.91 | 0.82 (0.08, 7.98) | 0.87 | 0.69 (0.07, 6.67) | 0.75 |
|  | 2017 | 447 | 9.03 | 0.74 (0.08, 7.15) | 0.79 | 0.56 (0.06, 5.43) | 0.62 |
|  | 2018 | 596 | 12.04 | 0.66 (0.07, 6.37) | 0.72 | 0.56 (0.06, 5.46) | 0.62 |
|  | 2019 | 349 | 7.05 | 0.60 (0.06, 5.85) | 0.66 | 0.48 (0.05, 4.69) | 0.53 |
|  | 2020 | 268 | 5.41 | 0.98 (0.1, 9.58) | 0.99 | 0.83 (0.08, 8.12) | 0.87 |
|  | 2021 | 29 | 0.59 | 0.95 (0.09, 10.71) | 0.97 | 0.95 (0.09, 10.71) | 0.97 |
|  | 2022 | 236 | 4.77 | 0.58 (0.06, 5.68) | 0.64 | 0.47 (0.05, 4.66) | 0.52 |

When considering only the 1,356 *emm*1 isolates that were assigned to a lineage, the 30-day CFR was 22.3% (118/530, 95% CI: 18.8%, 26.0%) for iGAS caused by M1_global_ and 24.4% (190/778, 95% CI: 21.4%, 27.6%) for iGAS caused by M1_UK_, a difference that was not significant. Interestingly, the 30-day CFR was much lower for iGAS caused by the intermediate lineages (M1_13SNP_: 10.3%, M1_23SNP_: 10.5%) although the denominators in these groups were lower (3/29 and 2/19 deaths respectively) (Figure 1). The 7-day case fatality rates followed a similar pattern (Supplementary Material Table S1).

**Figure 1.**
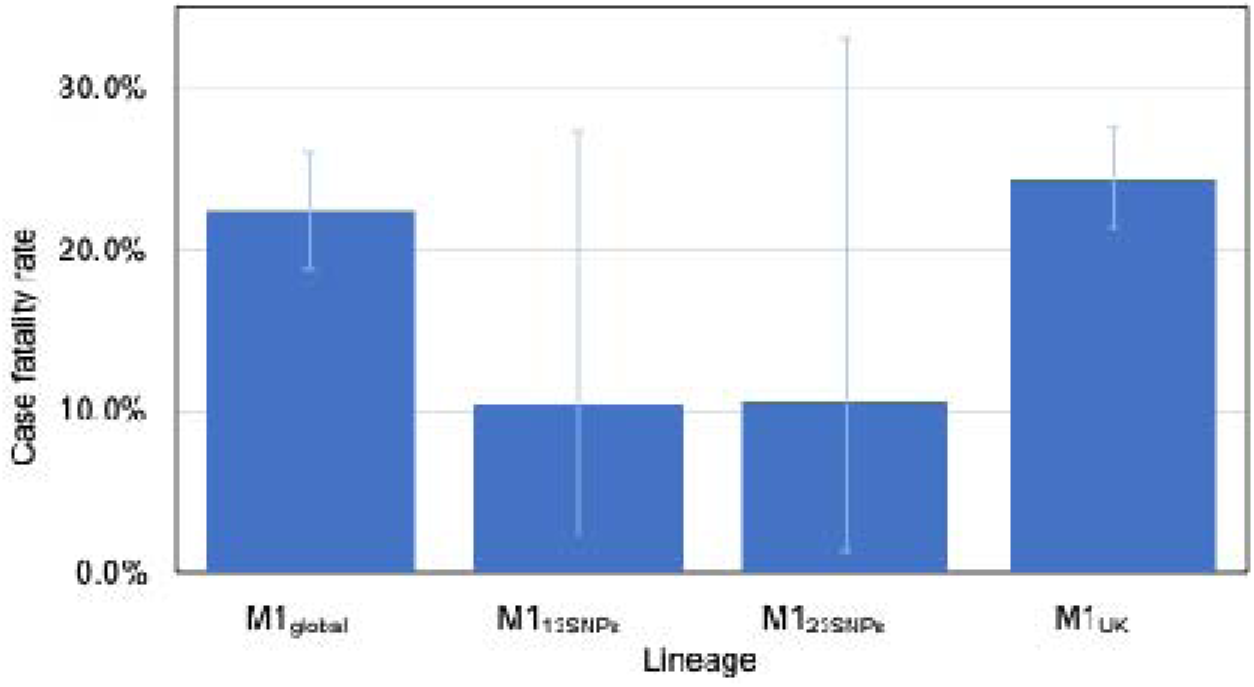
30-day case fatality rate by *emm*1 lineage. Columns show 30-day case fatality rate, error bars represent 95% CI. Data are from 1356 patients for whom lineage was assigned and with follow up data. Denominators for each group are M1_global_ (n=530); M1_13SNPs_ (n=29); M1_23SNPs_ (n=19); M1_UK_ (n=778).

### Survival analysis and time to death

Kaplan-Meier analysis comparing survival between M1_global_ and M1_UK_ iGAS cases over 30 days (Figure 2) revealed that the observed differences in survival were not significant (p = 0.24). Reduced survival observed for M1_UK_ iGAS cases was mostly ascribed to patients in the largest, 15-64 year old age group however differences in survival between M1_global_ and M1_UK_ iGAS were not significant for any age group (Supplementary Material Figure S2). Of note, almost all deaths beyond 3 days of sample were seen in the oldest age groups only.

**Figure 2.**
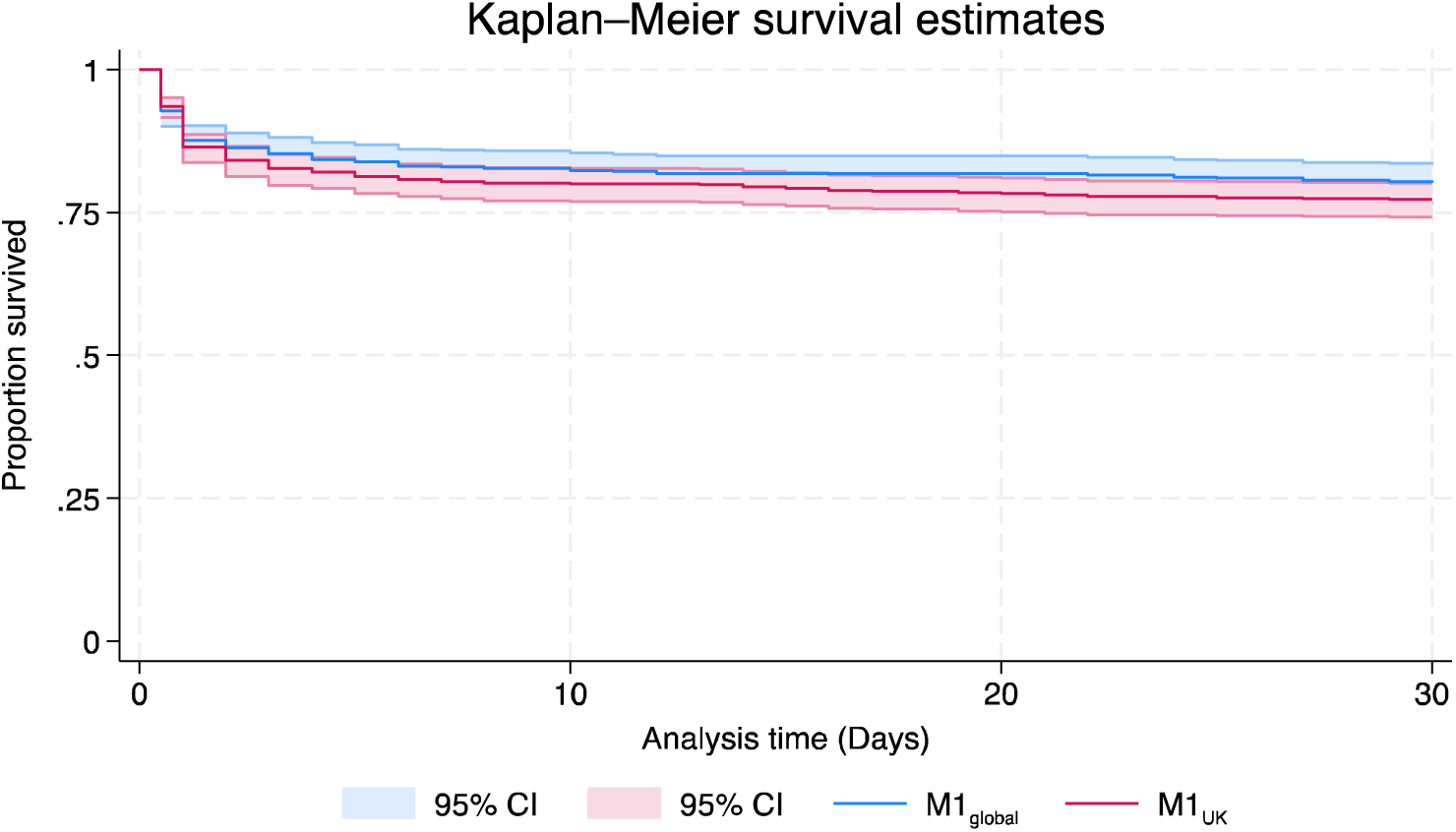
30-day survival of M1_global_ and M1_UK_ iGAS patients. Shaded regions show 95% CI. Number in each group M1_global_ (n=530); M1_UK_ (n=778). Those who died [day −7 to day-1] inclusive were recorded as dying at time zero 0. Those who died on day of sample were recorded as dying at time 0.5 days.

Considering all deaths from *emm*1 *S. pyogenes* we analysed the time from diagnostic sample being obtained to death (using an extended period, from −30 days to + 30 days from date of sample, n=1060 deaths, one of unknown age). Strikingly, among children under 15 years who died, a majority (56.3%, 76/135) died before any sample was taken, 87.4% (118/135) had died by the day of sampling, and 95.6% (129/135) had died by the day after sampling; only 2 deaths in children were observed after 2 days of sample being taken (Figure 3). Even among older age groups, almost all deaths had occurred by one day after a sample had been taken for culture (15y to 64y: 65%, 182/280; 65y to 84y: 57.5%, 226/393; 85y and above: 55%, 138/251) (Figure 3). When considering those deaths where *emm*1 isolates were assigned to a lineage, no difference between M1_global_ and M1_UK_ with regard to time to death was observed (Supplementary Figure S3).

**Figure 3.**
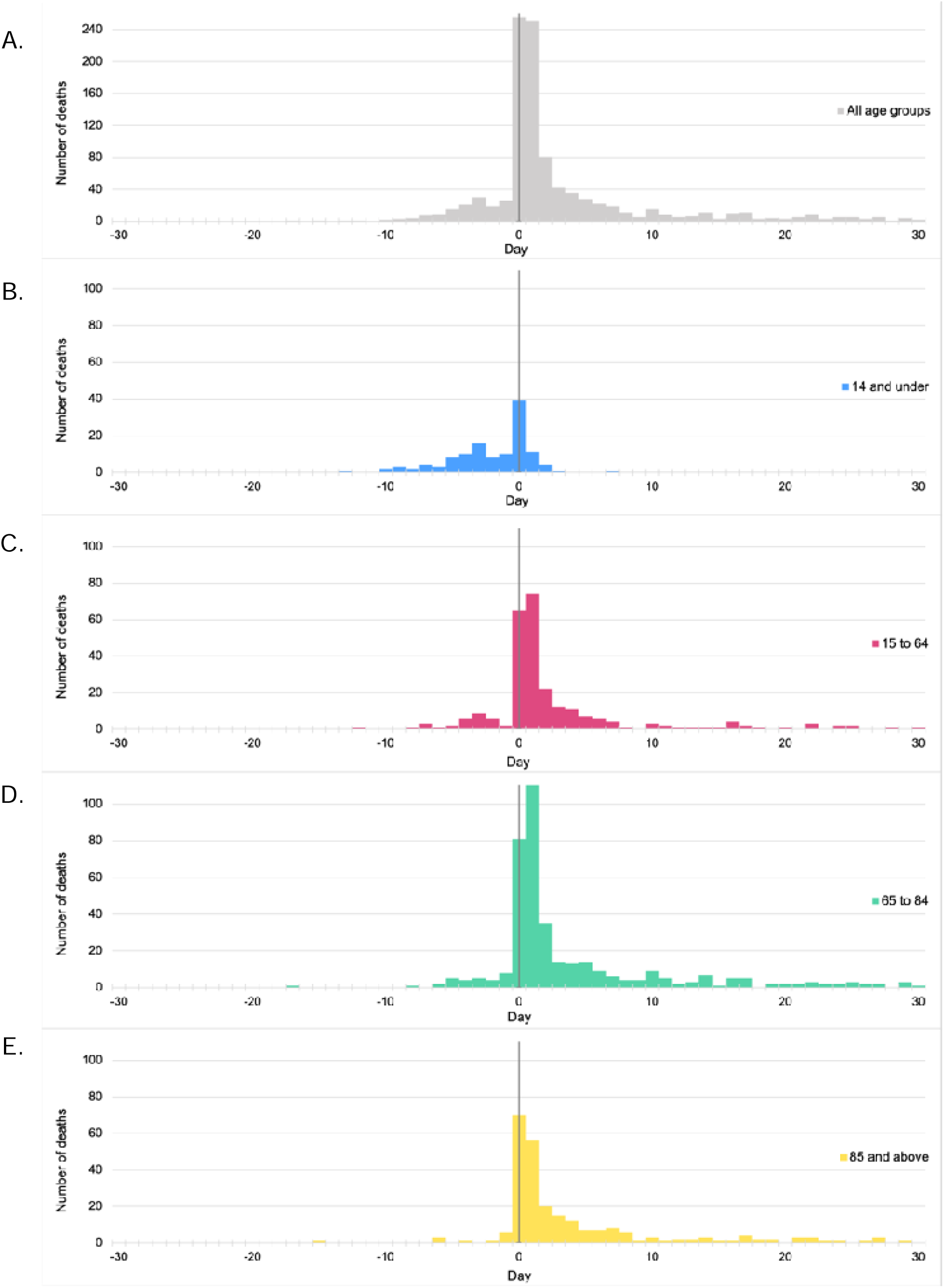
Time between sample collection and death by age. Time between diagnostic sample and death in different age groups. Each panel shows deaths from *emm*1 iGAS in all age groups (A, grey); in children aged 14y and under (B, blue); in adults aged 15y-64y (C, red); in adults aged 65y −84y (D, green); and in adults aged 85y and over (E, yellow). Y axes show numbers of deaths for each sublineage. X axes show number of days since date of diagnostic sample that yielded *S. pyogenes*.

### Univariate and multivariate analysis

Univariate analysis of all *emm1* cases indicated an increase in risk of 30-day mortality from iGAS among females compared with males (OR 1.16, 95% CI: 1.01-1.33, p, 0.04). Age had a greater impact on mortality, with those aged 65-84 having a 2-fold, and those aged 85 and older having a 6.5-fold increased risk of death compared with children (Table 1). Indeed, every year of age added an incremental 2% risk of death in adults. Although no effect of ethnicity or deprivation was observed overall, analysis by age subgroup pointed to an increased risk of mortality among Black and Asian children (under 15 years), and, to a non-significant extent, those of mixed ethnicity compared to white ethnicity (Supplementary Table S2).

To determine whether M1_UK_ was associated with increased risk of death, multivariate analysis was performed for the subcohort with complete data for lineage, and adjusted for age and sex (Table 2). After adjusting for these factors, age was the only factor conferring an increased risk of 30-day and 7-day mortality. Considering the cohort of iGAS patients with complete data, *emm*1 lineage did not have a significant effect.

**Table 2.**
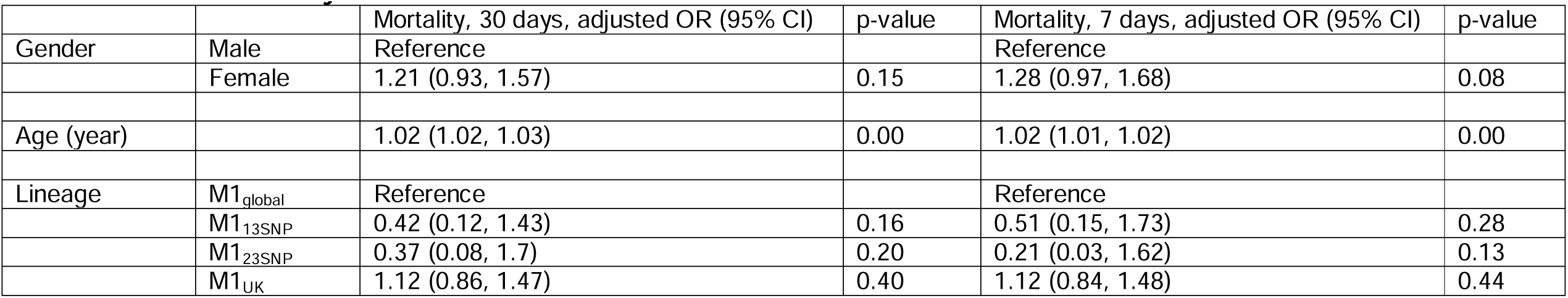
Multivariate analysis.

|  |  | Mortality, 30 days, adjusted OR (95% CI) | p-value | Mortality, 7 days, adjusted OR (95% CI) | p-value |
| --- | --- | --- | --- | --- | --- |
| Gender | Male | Reference |  | Reference |  |
|  | Female | 1.21 (0.93, 1.57) | 0.15 | 1.28 (0.97, 1.68) | 0.08 |
| Age (year) |  | 1.02 (1.02, 1.03) | 0.00 | 1.02 (1.01, 1.02) | 0.00 |
| Lineage | M1<sub>global</sub> | Reference |  | Reference |  |
|  | M1<sub>13SNP</sub> | 0.42 (0.12, 1.43) | 0.16 | 0.51 (0.15, 1.73) | 0.28 |
|  | M1<sub>23SNP</sub> | 0.37 (0.08, 1.7) | 0.20 | 0.21 (0.03, 1.62) | 0.13 |
|  | M1<sub>UK</sub> | 1.12 (0.86, 1.47) | 0.40 | 1.12 (0.84, 1.48) | 0.44 |

## Discussion

We set out to determine the attributable case fatality of iGAS caused by the emergent lineage M1_UK_ compared with ancestral *emm*1 strains. We found the average case fatality rate (CFR) associated with M1_UK_ iGAS in the pre-COVID19 pandemic period to be 24.4%, which is higher than the reported CFR for iGAS overall in the USA (1), but not significantly different to the CFR for iGAS caused by M1_global_ isolates (22.3%). The very high CFR associated with *emm*1 *S. pyogenes* was notably more than double the CFR measured for the intermediate *emm*1 lineages.

A striking observation was the rapidity of death caused by *emm*1 iGAS infections regardless of lineage, a feature of iGAS that has been highlighted before (24). A majority of deaths occurred within just one day of a sample being taken for culture, the earliest point that culture-based diagnosis might be obtained. Indeed, in children, >50% deaths occurred before any sample was taken, underlining a need for case-based reviews to identify what interventions might improve outcome, including implementation of rapid diagnostic tests, value of clinical severity scores, and merits of enhancing prevention of transmission during outbreaks. The rapidity of deaths has implications for clinical trial design: for *emm*1 iGAS, waiting for culture-based diagnostic confirmation may make any intervention futile as any deaths attributable to *S. pyogenes* will already have happened. Few bacterial infections progress as rapidly as iGAS in previously healthy people, contributing to an acute sense of injustice among the bereaved when deaths occur. Although we did not identify social deprivation as a risk factor, univariate analysis indicated non-white ethnicity in children to be associated with increased CFR; whether this is linked to healthcare access will be examined in future studies.

Although M1_UK_ was not significantly more lethal than M1_global_ in the cohort examined, enhanced fitness for transmission is believed to underlie its increasing dominance. Whether this is linked to the increased production of superantigen SpeA or other metabolic attributes is unclear (38). A number of studies report that M1_UK_ isolates display minimal genetic change over a fixed time period, indicating genomic stability despite widespread transmission (7,16,18) with dominance even in non-invasive infections (39). As population immunity evolves, it may be that any advantage dwindles. Given that M1_UK_ dominated many of the reported iGAS upsurges in the post-pandemic period (6–8, 14,16), our findings go some way to explaining the observed numbers of fatalities in such upsurges (14, 23). The markedly lower CFR (10%) observed following iGAS with intermediate lineages M1_13SNP_ and M1_23SNP_ was surprising, albeit that these groups comprised just 48 patients, making statistical comparison challenging. The data suggest that these intermediate lineages do not share the fitness to cause severe invasive infection that is seen in the major dominant *emm*1 lineages, despite M1_23SNP_ isolates expressing SpeA to the same level as M1_UK_ strains (38). Arising around 2002 and 2006 respectively as antecedents of M1_UK_, these intermediate lineages are no longer detectable in England. (16, 39)

Our study was limited by an inability to balance cases from each lineage over time, due to the very rapid expansion of M1_UK_ in England between 2010 and 2015 (18), and by the number of iGAS cases for which lineage had been assigned. Although we deliberately included cases from 2010, the year iGAS became statutorily notifiable in England, to increase the cohort size with M1_global_ infection, linkage to outcome data was not as successful as more recent years. Given the CFR difference observed, the study lacked power to demonstrate a significant difference between the two major *emm*1 lineages. An ideal study might require >9,000 cases in each group, which is not feasible even if including all cases of *emm*1 iGAS in England and Wales. Formal comparison of CFR would be valuable from other regions where both lineages currently co-exist, using agreed standard definitions to allow meta-analysis. Active surveillance from CDC from ten US states 2015-2021 reported a CFR of 22% for M1_UK_ and 14.9% for M1_global_ however this did not reach significance, likely due to a 20-fold imbalance between lineage-associated cohort size (40).

Although we were able to include ethnicity and markers of deprivation in our analysis, we were unable to include comorbidity, which can have a major influence on outcome in many bacterial infections including iGAS. As with all retrospective analyses, it is possible that iGAS outcome more generally may have improved over time due to changes in sepsis management, for example more widespread use of clindamycin, potentially impacting results. Despite this, the CFR of M1_UK_ was somewhat greater than M1_global_ in all age groups bar the over-85s, albeit not significantly. Our study is unlikely to have been affected by COVID-19, as the vast majority of *emm*1 iGAS in 2020 was in Q1 only, after which *emm*1 iGAS was rare. We also excluded the post-COVID-19 iGAS upsurge period, to avoid the impact of any population immunity gap and respiratory viruses on outcomes (15,16).

Emergence of a new variant of any pathogen should prompt rapid evaluation of the potential for harm; while this is well-understood for viruses of pandemic potential, the principle is applicable to bacteria and other microorganisms as well. This is true not only for bacterial lineages that harbour specific antimicrobial resistance traits, but acutely relevant for bacteria of high pathogenicity such as *S. pyogenes*, for which there are no vaccines. For rare infections this may require multi-country meta-analysis. As an exclusively human pathogen, better understanding of the severity that can be attributed to different lineages will be of great value in planning for, and managing, the public’s health.

## Data Availability

All de-identified data produced in the present study are available upon reasonable request to the authors

## Acknowledgements

The authors are grateful to the diagnostic bacteriology laboratories throughout England and Wales that submit isolates to the Reference laboratory, and to the Reference laboratory for *emm* typing isolates. They also acknowledge support provided by the NIHR Imperial Biomedical Research Centre awarded to Imperial College London and the Conor Kerin Foundation.

## Author contributions

Project conceptualisation, SS, TL, HKL; Data acquisition, HKL, OW, RD, JC; Data linkage RG; TL; Data curation RG, NZ; Data analysis HKL, NZ; Data visualisation NZ; Manuscript original draft SS, HKL, NZ; Manuscript review and editing, all authors; Final approval, all authors

## References

1. Gregory CJ, Okaro JO, Reingold A, Chai S, Herlihy R, Petit S, Farley MM, Harrison LH, Como-Sabetti K, Lynfield R, Snippes Vagnone P, Sosin D, Anderson BJ, Burzlaff K, Martin T, Thomas A, Schaffner W, Talbot HK, Beall B, Chochua S, Chung Y, Park S, Van Beneden C, Li Y, Schrag SJ. Invasive Group A Streptococcal Infections in 10 US States. JAMA. 2025 May 6;333(17):1498–1507. doi: 10.1001/jama.2025.0910.

2. Luca-Harari B, Darenberg J, Neal S, Siljander T, Strakova L, Tanna A, Creti R, Ekelund K, Koliou M, Tassios PT, van der Linden M, Straut M, Vuopio-Varkila J, Bouvet A, Efstratiou A, Schalén C, Henriques-Normark B; Strep-EURO Study Group; Jasir A. Clinical and microbiological characteristics of severe Streptococcus pyogenes disease in Europe. J Clin Microbiol. 2009 Apr;47(4):1155–65. doi: 10.1128/JCM.02155-08

3. WHO. Increased incidence of scarlet fever and invasive Group A Streptococcus infection - multi-country. December 2022. https://www.who.int/emergencies/disease-outbreak-news/item/2022-DON429

4. Pan American Health Organization / World Health Organization. Epidemiological alert:Invasive disease caused by group A streptococci - 28 November 2023. Washington, D.C. https://www.paho.org/en/documents/epidemiological-alert-invasive-disease-caused-group-streptococci-28-november-2023

5. Guy R, Henderson KL, Coelho J, Hughes H, Mason EL, Gerver SM, Demirjian A, Watson C, Sharp A, Brown CS, Lamagni T. Increase in invasive group A streptococcal infection notifications, England, 2022. Euro Surveill. 2023 Jan;28(1):2200942. doi: 10.2807/1560-7917.ES.2023.28.1.2200942

6. Rümke LW, Davies MA, Vestjens SMT, van der Putten BCL, Bril-Keijzers WCM, van Houten MA, Rots NY, Wijmenga-Monsuur AJ, van der Ende A, de Gier B, Vlaminckx BJM, van Sorge NM. Nationwide upsurge in invasive disease in the context of longitudinal surveillance of carriage and invasive *Streptococcus pyogenes* 2009-2023, the Netherlands: a molecular epidemiological study. J Clin Microbiol. 2024 Oct 16;62(10):e0076624. doi: 10.1128/jcm.00766-24.

7. Rodriguez-Ruiz JP, Lin Q, Lammens C, Smeesters PR, van Kleef-van Koeveringe S, Matheeussen V, Malhotra-Kumar S. Increase in bloodstream infections caused by *emm*1 group A *Streptococcus* correlates with emergence of toxigenic M1_UK_, Belgium, May 2022 to August 2023. Euro Surveill. 2023 Sep;28(36):2300422. doi: 10.2807/1560-7917.ES.2023.28.36.2300422.

8. Gouveia C, Bajanca-Lavado MP, Mamede R, Araújo Carvalho A, Rodrigues F, Melo-Cristino J, Ramirez M, Friães A; Portuguese Group for the Study of Streptococcal Infections; Portuguese Study Group of Pediatric Invasive Streptococcal Disease; Portuguese Study Group of Paediatric Invasive Streptococcal Disease. Sustained increase of paediatric invasive *Streptococcus pyogenes* infections dominated by M1_UK_ and diverse *emm*12 isolates, Portugal, September 2022 to May 2023. Euro Surveill. 2023 Sep;28(36):2300427. doi: 10.2807/1560-7917.ES.2023.28.36.2300427.

9. Johannesen TB, Munkstrup C, Edslev SM, Baig S, Nielsen S, Funk T, Kristensen DK, Jacobsen LH, Ravn SF, Bindslev N, Gubbels S, Voldstedlund M, Jokelainen P, Hallstrøm S, Rasmussen A, Kristinsson KG, Fuglsang-Damgaard D, Dessau RB, Olsén AB, Jensen CS, Skovby A, Ellermann-Eriksen S, Jensen TG, Dzajic E, Østergaard C, Lomborg Andersen S, Hoffmann S, Andersen PH, Stegger M. Increase in invasive group A streptococcal infections and emergence of novel, rapidly expanding sub-lineage of the virulent *Streptococcus pyogenes* M1 clone, Denmark, 2023. Euro Surveill. 2023 Jun;28(26):2300291. doi: 10.2807/1560-7917.ES.2023.28.26.2300291.

10. Ramírez de Arellano E, Saavedra-Lozano J, Villalón P, Jové-Blanco A, Grandioso D, Sotelo J, Gamell A, González-López JJ, Cervantes E, Gónzalez MJ, Rello-Saltor V, Esteva C, Sanz-Santaeufemia F, Yagüe G, Manzanares Á, Brañas P, Ruiz de Gopegui E, Carrasco-Colom J, García F, Cercenado E, Mellado I, Del Castillo E, Pérez-Vazquez M, Oteo-Iglesias J, Calvo C; Spanish PedGAS-Net/CIBERINFEC GAS Study Group. Clinical, microbiological, and molecular characterization of pediatric invasive infections by Streptococcus pyogenes in Spain in a context of global outbreak. mSphere. 2024 Mar 26;9(3):e0072923. doi: 10.1128/msphere.00729-23.

11. Golden AR, Griffith A, Tyrrell GJ, Kus JV, McGeer A, Domingo MC, Grant J, Minion J, Van Caeseele P, Desnoyers G, Haldane D, Yu Y, Ding X, Steven L, McFadzen J, Primeau C, Martin I. Invasive Group A Streptococcus Hypervirulent M1_UK_ Clone, Canada, 2018-2023. Emerg Infect Dis. 2024 Nov;30(11):2409–2413. doi: 10.3201/eid3011.241068.

12. Abo YN, Oliver J, McMinn A, Osowicki J, Baker C, Clark JE, Blyth CC, Francis JR, Carr J, Smeesters PR, Crawford NW, Steer AC. Increase in invasive group A streptococcal disease among Australian children coinciding with northern hemisphere surges. Lancet Reg Health West Pac. 2023 Aug 22;41:100873. doi: 10.1016/j.lanwpc.2023.100873.

13. Ammar S, Anglemyer A, Bennett J, Lees J, Addidle M, Morgan J, DuBray K, Galloway Y, Grey C, Duff P. Post-pandemic increase in invasive group A strep infections in New Zealand. J Infect Public Health. 2024 Nov;17(11):102545. doi: 10.1016/j.jiph.2024.102545.

14. National Institute for Infectious Diseases, July 2024 Risk Assessment for Streptococcal Toxic Shock Syndrome (STSS) in Japan https://www.niid.go.jp/niid/images/cepr/RA/STSS/240701_NIID_STSS_2_Eng.pdf

15. Messacar K, Baker RE, Park SW, Nguyen-Tran H, Cataldi JR, Grenfell B. Preparing for uncertainty: endemic paediatric viral illnesses after COVID-19 pandemic disruption. Lancet. 2022 Nov 12;400(10364):1663–1665. doi: 10.1016/S0140-6736(22)01277-6

16. Vieira A, Wan Y, Ryan Y, Li HK, Guy RL, Papangeli M, Huse KK, Reeves LC, Soo VWC, Daniel R, Harley A, Broughton K, Dhami C, Ganner M, Ganner MA, Mumin Z, Razaei M, Rundberg E, Mammadov R, Mills EA, Sgro V, Mok KY, Didelot X, Croucher NJ, Jauneikaite E, Lamagni T, Brown CS, Coelho J, Sriskandan S. Rapid expansion and international spread of M1_UK_ in the post-pandemic UK upsurge of Streptococcus pyogenes. Nat Commun. 2024 May 10;15(1):3916. doi: 10.1038/s41467-024-47929-7.

17. Vesty A, Ren X, Sharma P, Lorenz N, Proft T, Hardaker A, Straub C, Morgan J, Tiong A, Anderson A, Webb RH, Bennett J, Carter PE, Moreland NJ. The Emergence and Impact of the M1_UK_ Lineage on Invasive Group A Streptococcus Disease in Aotearoa New Zealand. Open Forum Infect Dis. 2024 Aug 9;11(8):ofae457. doi: 10.1093/ofid/ofae457.

18. Lynskey NN, Jauneikaite E, Li HK, Zhi X, Turner CE, Mosavie M, Pearson M, Asai M, Lobkowicz L, Chow JY, Parkhill J, Lamagni T, Chalker VJ, Sriskandan S. Emergence of dominant toxigenic M1T1 Streptococcus pyogenes clone during increased scarlet fever activity in England: a population-based molecular epidemiological study. Lancet Infect Dis. 2019 Nov;19(11):1209–1218. doi: 10.1016/S1473-3099(19)30446-3.

19. Bertram R, Itzek A, Marr L, Manzke J, Voigt S, Chapot V, van der Linden M, Rath P-M, Hitzl W, Steinmann J. Divergent effects of *emm* types 1 and 12 on invasive group A streptococcal infections-results of a retrospective cohort study, Germany 2023. J Clin Microbiol. 2024 Aug 14;62(8):e0063724. doi: 10.1128/jcm.00637-24 (70% are M1UK)Found not more invasive references dansih paper also?

20. Veselá R, Vohrnová S, Kozáková J. Výskyt sub-linie M1 UK mezi invazivními kmeny Streptococcus pyogenes typ emm1 izolovanými od prosince 2022 do května 2023 v České republice [Prevalence of the M1UK sublineage among emm1 Streptococcus pyogenes invasive strains isolated in the Czech Republic from December 2022 to May 2023]. Epidemiol Mikrobiol Imunol. 2024;73(2):76–83. Czech. doi: 10.61568/emi/11-6306/20240424/137080.

21. Davies MR, Keller N, Brouwer S, Jespersen MG, Cork AJ, Hayes AJ, Pitt ME, De Oliveira DMP, Harbison-Price N, Bertolla OM, Mediati DG, Curren BF, Taiaroa G, Lacey JA, Smith HV, Fang NX, Coin LJM, Stevens K, Tong SYC, Sanderson-Smith M, Tree JJ, Irwin AD, Grimwood K, Howden BP, Jennison AV, Walker MJ. Detection of Streptococcus pyogenes M1_UK_ in Australia and characterization of the mutation driving enhanced expression of superantigen SpeA. Nat Commun. 2023 Feb 24;14(1):1051. doi: 10.1038/s41467-023-36717-4.

22. Lees EA, Williams TC, Marlow R, Fitzgerald F, Jones C, Lyall H, Bamford A, Pollock L, Smith A, Lamagni T, Kent A, Whittaker E; Group A Streptococcal Disease Consortium. Epidemiology and Management of Pediatric Group A Streptococcal Pneumonia With Parapneumonic Effusion: An Observational Study. Pediatr Infect Dis J. 2024 Sep 1;43(9):841–850. doi: 10.1097/INF.0000000000004418.

23. Wrenn K, Blomquist PB, Inzoungou-Massanga C, Olufon O, Guy RL, Hatziioanou D, Findlater L, Smith I, Mirfenderesky M, Luyt K, Williams T, Stoianova S, Dickinson M, Pietzsch M, Jarvis CI, Brown C, Lamagni T, Kumar D. Surge of lower respiratory tract group A streptococcal infections in England in winter 2022: epidemiology and clinical profile. Lancet. 2023 Nov;402 Suppl 1:S93. doi: 10.1016/S0140-6736(23)02095-0.

24. Lamagni TL, Neal S, Keshishian C, Powell D, Potz N, Pebody R, George R, Duckworth G, Vuopio-Varkila J, Efstratiou A. Predictors of death after severe Streptococcus pyogenes infection. Emerg Infect Dis. 2009 Aug;15(8):1304–7. doi: 10.3201/eid1508.090264

25. Sumby P, Whitney AR, Graviss EA, DeLeo FR, Musser JM. Genome-wide analysis of group a streptococci reveals a mutation that modulates global phenotype and disease specificity. PLoS Pathog. 2006 Jan;2(1):e5. doi: 10.1371/journal.ppat.0020005

26. Friães A, Pato C, Melo-Cristino J, Ramirez M. Consequences of the variability of the CovRS and RopB regulators among Streptococcus pyogenes causing human infections. Sci Rep. 2015 Jul 15;5:12057. doi: 10.1038/srep12057

27. Bergsten H, Nizet V. The intricate pathogenicity of Group A *Streptococcus*: A comprehensive update. Virulence. 2024 Dec;15(1):2412745. doi: 10.1080/21505594.2024.2412745

28. Sumby P, Porcella SF, Madrigal AG, Barbian KD, Virtaneva K, Ricklefs SM, Sturdevant DE, Graham MR, Vuopio-Varkila J, Hoe NP, Musser JM. Evolutionary origin and emergence of a highly successful clone of serotype M1 group A Streptococcus involved multiple horizontal gene transfer events. J Infect Dis. 2005 Sep 1;192(5):771–82. doi: 10.1086/432514

29. Li Y, Nanduri SA, Van Beneden CA, Beall BW. M1_UK_ lineage in invasive group A streptococcus isolates from the USA. Lancet Infect Dis. 2020 May;20(5):538–539. doi: 10.1016/S1473-3099(20)30279-6.

30. NHS England. The NHS Spine. 2024. https://digital.nhs.uk/services/spine.

31. NHS England. The Personal Demographics Service (PDS). 2024. Personal Demographics Service - NHS England Digital

32. NHS England. Hospital Episode Statistics (HES). 2023. https://digital.nhs.uk/data-and-information/data-tools-and-services/dataservices/hospital-episode-statistics

33. Office for Health Improvement & Disparities. Method for assigning ethnic group in the COVID-19 Health Inequalities Monitoring for England (CHIME) tool. 2022. https://www.gov.uk/government/statistics/covid-19-health-inequalities-monitoring-in-england-tool-chime/method-for-assigning-ethnic-group-in-the-covid-19-health-inequalities-monitoring-for-england-chime-tool

34. Zhi X, Li HK, Li H, Loboda Z, Charles S, Vieira A, Huse K, Jauneikaite E, Reeves L, Mok KY, Coelho J, Lamagni T, Sriskandan S. Emerging Invasive Group A Streptococcus M1_UK_ Lineage Detected by Allele-Specific PCR, England, 2020^1^. Emerg Infect Dis. 2023 May;29(5):1007–1010. doi: 10.3201/eid2905.221887

35. N. Dudley, P. W., Wickham, P. R. A. R., & Coombs, M. N. (2016). An Introduction to Survival Statistics: Kaplan-Meier Analysis. Journal of the Advanced Practitioner in Oncology, 7(1). 10.6004/jadpro.2016.7.1.8

36. Bewick, V., Cheek, L., & Ball, J. (2004). Statistics review 12: Survival analysis. Critical Care, 8(5), 389. 10.1186/cc2955

37. Colosimo, E., Ferreira, F., Oliveira, M., & Sousa, C. (2002). Empirical comparisons between Kaplan-Meier and Nelson-Aalen survival function estimators. Journal of Statistical Computation and Simulation, 72(4), 299–308. 10.1080/00949650212847

38. Li HK, Zhi X, Vieira A, Whitwell HJ, Schricker A, Jauneikaite E, Li H, Yosef A, Andrew I, Game L, Turner CE, Lamagni T, Coelho J, Sriskandan S. Characterization of emergent toxigenic M1_UK_*Streptococcus pyogenes* and associated sublineages. Microb Genom. 2023 Apr;9(4):mgen000994. doi: 10.1099/mgen.0.000994

39. Hall JN, Bah SY, Khalid H, Brailey A, Coleman S, Kirk T, Hussain N, Tovey M, Chaudhuri RR, Davies S, Tilley L, de Silva T, Turner CE. Molecular characterization of *Streptococcus pyogenes* (StrepA) non-invasive isolates during the 2022-2023 UK upsurge. Microb Genom. 2024 Aug;10(8):001277. doi: 10.1099/mgen.0.001277.

40. Li Y, Rivers J, Mathis S, et al. Expansion of Invasive Group A Streptococcus M1UK Lineage in Active Bacterial Core Surveillance, United States, 2019=2021. Emerging Infectious Diseases. 2023;29(10):2116-2120. doi:10.3201/eid2910.230675

