## Supplementary tables and figures for "Mortality among patients with invasive group A streptococcal infections caused by the M1_UK_ lineage: a retrospective cohort study in England and Wales"

**Li HK, Zhu N, et al.**

**Supplementary Tables**

Table S1 Case fatality rate by age group. Page 2

Table S2 Univariate analysis by age group Page 3

**Supplementary Figures**

Figure S1 *emm*1 iGAS cases per month and proportion assigned to lineage Page 7

Figure S2 30-day survival of $M1_{global}$ and $M1_{UK}$ iGAS cases by age group Page 8

Figure S3 Time between sample and death by lineage Page 9

**Table S1 Case fatality rate by age group**

| All age groups | | | | | |
|---|---|---|---|---|---|
| | **All** | **M1**$_{global}$ | **M1**$_{13SNP}$ | **M1**$_{23SNP}$ | **M1**$_{UK}$ |
| Total | 4,952 | 530 | 29 | 19 | 778 |
| Died, 30 days | 1,043 | 118 | 3 | 2 | 190 |
| CFR, 30 days (95% CI) | 0.211 [0.199, 0.222] | 0.223 [0.188, 0.260] | 0.103 [0.022, 0.274] | 0.105 [0.013, 0.331] | 0.244 [0.214, 0.276] |
| Died, 7 days | 901 | 104 | 3 | 1 | 167 |
| CFR, 7 days (95% CI) | 0.182 [0.171, 0.193] | 0.196 [0.163, 0.233] | 0.103 [0.022, 0.274] | 0.053 [0.001, 0.260] | 0.215 [0.186, 0.245] |
| Under 15 years | | | | | |
| | **All** | **M1**$_{global}$ | **M1**$_{13SNP}$ | **M1**$_{23SNP}$ | **M1**$_{UK}$ |
| Total | 950 | 100 | 5 | 5 | 149 |
| Died, 30 days | 124 | 17 | 0 | 0 | 19 |
| CFR, 30 days (95% CI) | 0.131 [0.110, 0.154] | 0.170 [0.102, 0.258] | 0.000 [0.000, 0.522] | 0.000 [0.000, 0.522] | 0.128 [0.079, 0.192] |
| Died, 7 days | 124 | 17 | 0 | 0 | 19 |
| CFR, 7 days (95% CI) | 0.131 [0.110, 0.154] | 0.170 [0.102, 0.258] | 0.000 [0.000, 0.522] | 0.000 [0.000, 0.522] | 0.128 [0.079, 0.192] |
| 15 to 64 years | | | | | |
| | **All** | **M1**$_{global}$ | **M1**$_{13SNP}$ | **M1**$_{23SNP}$ | **M1**$_{UK}$ |
| Total | 1887 | 200 | 12 | 5 | 296 |
| Died, 30 days | 277 | 26 | 0 | 0 | 52 |
| CFR, 30 days (95% CI) | 0.147 [0.131, 0.164] | 0.130 [0.087, 0.185] | 0.000 [0.000, 0.265] | 0.000 [0.000, 0.522] | 0.176 [0.134, 0.224] |
| Died, 7 days | 248 | 22 | 0 | 0 | 50 |
| CFR, 7 days (95% CI) | 0.131 [0.116, 0.148] | 0.110 [0.070, 0.162] | 0.000 [0.000, 0.265] | 0.000 [0.000, 0.522] | 0.169 [0.128, 0.217] |
| 65 to 84 years | | | | | |
| | **All** | **M1**$_{global}$ | **M1**$_{13SNP}$ | **M1**$_{23SNP}$ | **M1**$_{UK}$ |
| Total | 1606 | 184 | 10 | 5 | 239 |
| Died, 30 days | 391 | 50 | 2 | 0 | 72 |
| CFR, 30 days (95% CI) | 0.243 [0.223, 0.265] | 0.272 [0.209, 0.342] | 0.200 [0.025, 0.556] | 0.000 [0.000, 0.522] | 0.234 [0.175, 0.302] |
| Died, 7 days | 320 | 43 | 2 | 0 | 63 |
| CFR, 7 days (95% CI) | 0.199 [0.180, 0.220] | 0.301 [0.244, 0.364] | 0.200 [0.025, 0.556] | 0.000 [0.000, 0.522] | 0.264 [0.209, 0.324] |
| 85 years and above | | | | | |
| | **All** | **M1**$_{global}$ | **M1**$_{13SNP}$ | **M1**$_{23SNP}$ | **M1**$_{UK}$ |
| Total | 505 | 46 | 2 | 4 | 93 |
| Died, 30 days | 250 | 25 | 1 | 2 | 47 |
| CFR, 30 days (95% CI) | 0.495 [0.451, 0.540] | 0.543 [0.390, 0.691] | 0.500 [0.013, 0.987] | 0.500 [0.068, 0.932] | 0.478 [0.329, 0.631] |
| Died, 7 days | 208 | 22 | 1 | 1 | 35 |
| CFR, 7 days (95% CI) | 0.412 [0.369, 0.456] | 0.505 [0.400, 0.611] | 0.500 [0.013, 0.987] | 0.250 [0.006, 0.806] | 0.376 [0.278, 0.483] |

**Table S2 Univariate analysis by age group**

| | | Number (N) | Percentage (%) | Mortality, 30 days, odds ratio (95% CI) | p-value | Mortality, 7 days, odds ratio (95% CI) | p-value |
|---|---|---|---|---|---|---|---|
| Under 15 | | | | | | | |
| Gender | Male | 500 | 52.63 | Reference | | Reference | |
| | Female | 440 | 46.32 | 1.08 (0.74, 1.57) | 0.71 | 1.08 (0.74, 1.57) | 0.71 |
| | Unknown | 10 | 1.05 | | | | |
| Ethnicity | White | 618 | 65.05 | Reference | | Reference | |
| | Asian | 114 | 12.00 | 1.76 (1.01, 3.08) | 0.05 | 1.76 (1.01, 3.08) | 0.05 |
| | Black | 52 | 5.47 | 2.94 (1.49, 5.79) | 0.00 | 2.94 (1.49, 5.79) | 0.00 |
| | Mixed | 36 | 3.79 | 2.13 (0.89, 5.05) | 0.09 | 2.13 (0.89, 5.05) | 0.09 |
| | Other | 9 | 0.95 | 2.52 (0.51, 12.38) | 0.26 | 2.52 (0.51, 12.38) | 0.26 |
| | Unknown | 121 | 12.74 | | | | |
| IMD | 1 | 206 | 21.68 | Reference | | Reference | |
| | 2 | 116 | 12.21 | 0.47 (0.21, 1.07) | 0.07 | 0.47 (0.21, 1.07) | 0.07 |
| | 3 | 116 | 12.21 | 1.25 (0.66, 2.34) | 0.50 | 1.25 (0.66, 2.34) | 0.50 |
| | 4 | 91 | 9.58 | 0.87 (0.41, 1.84) | 0.72 | 0.87 (0.41, 1.84) | 0.72 |
| | 5 | 76 | 8.00 | 0.54 (0.22, 1.37) | 0.20 | 0.54 (0.22, 1.37) | 0.20 |
| | Unknown | 345 | 36.32 | | | | |
| Lineage | $M1_{global}$ | 100 | 10.53 | Reference | | Reference | |
| | $M1_{13SNP}$ | 5 | 0.53 | Empty | | Empty | |
| | $M1_{23SNP}$ | 5 | 0.53 | Empty | | Empty | |
| | $M1_{UK}$ | 149 | 15.68 | 0.71 (0.35, 1.45) | 0.35 | 0.71 (0.35, 1.45) | 0.35 |
| | Unknown | 691 | 72.74 | | | | |
| Sample year | 2009 | 1 | 0.11 | Reference | | Reference | |
| | 2010 | 77 | 8.11 | 0.85 (0.32, 2.27) | 0.74 | 0.85 (0.32, 2.27) | 0.74 |
| | 2011 | 82 | 8.63 | 0.9 (0.35, 2.34) | 0.83 | 0.9 (0.35, 2.34) | 0.83 |
| | 2012 | 79 | 8.32 | 1.44 (0.59, 3.5) | 0.42 | 1.44 (0.59, 3.5) | 0.42 |
| | 2013 | 86 | 9.05 | 1.18 (0.48, 2.91) | 0.71 | 1.18 (0.48, 2.91) | 0.71 |
| | 2014 | 59 | 6.21 | 1.15 (0.42, 3.1) | 0.79 | 1.15 (0.42, 3.1) | 0.79 |
| | 2015 | 97 | 10.21 | 1.34 (0.57, 3.16) | 0.51 | 1.34 (0.57, 3.16) | 0.51 |
| | 2016 | 115 | 12.11 | 1.63 (0.72, 3.68) | 0.24 | 1.63 (0.72, 3.68) | 0.24 |
| | 2017 | 67 | 7.05 | 0.72 (0.25, 2.09) | 0.54 | 0.72 (0.25, 2.09) | 0.54 |
| | 2018 | 100 | 10.53 | 1 (0.41, 2.44) | 0.99 | 1 (0.41, 2.44) | 0.99 |
| | 2019 | 54 | 5.68 | 1.09 (0.39, 3.05) | 0.87 | 1.09 (0.39, 3.05) | 0.87 |
| | 2020 | 42 | 4.42 | 0.37 (0.08, 1.75) | 0.21 | 0.37 (0.08, 1.75) | 0.21 |
| | 2021 | 8 | 0.84 | 1.04 (0.12, 9.38) | 0.97 | 1.04 (0.12, 9.38) | 0.97 |

| | | | | | | | |
|---|---|---|---|---|---|---|---|
| | 2022 | 83 | 8.74 | Empty | | Empty | |
| 15 to 64 | | | | | | | |
| Gender | Male | 992 | 52.57 | Reference | | Reference | |
| | Female | 885 | 46.9 | 0.97 (0.75, 1.26) | 0.83 | 1.04 (0.79, 1.36) | 0.78 |
| | Unknown | 10 | 0.53 | | | | |
| Ethnicity | White | 1,453 | 77.00 | Reference | | Reference | |
| | Asian | 109 | 5.78 | 0.90 (0.5, 1.6) | 0.71 | 0.93 (0.51, 1.69) | 0.81 |
| | Black | 55 | 2.91 | 0.48 (0.17, 1.34) | 0.16 | 0.40 (0.12, 1.28) | 0.12 |
| | Mixed | 17 | 0.90 | 1.30 (0.37, 4.58) | 0.68 | 1.47 (0.42, 5.16) | 0.55 |
| | Other | 20 | 1.06 | 1.52 (0.5, 4.6) | 0.46 | 1.71 (0.57, 5.18) | 0.34 |
| | Unknown | 233 | 12.35 | | | | |
| IMD | 1 | 296 | 15.69 | Reference | | Reference | |
| | 2 | 258 | 13.67 | 0.98 (0.63, 1.54) | 0.94 | 1.11 (0.7, 1.77) | 0.66 |
| | 3 | 257 | 13.62 | 0.63 (0.38, 1.02) | 0.06 | 0.52 (0.3, 0.91) | 0.02 |
| | 4 | 258 | 13.67 | 0.72 (0.45, 1.16) | 0.18 | 0.83 (0.51, 1.36) | 0.47 |
| | 5 | 215 | 11.39 | 0.80 (0.49, 1.30) | 0.37 | 0.95 (0.58, 1.58) | 0.86 |
| | Unknown | 603 | 31.96 | | | | |
| Lineage | $M1_{global}$ | 200 | 10.60 | Reference | | Reference | |
| | $M1_{13SNP}$ | 12 | 0.64 | Empty | | Empty | |
| | $M1_{23SNP}$ | 5 | 0.26 | Empty | | Empty | |
| | $M1_{UK}$ | 296 | 15.69 | 1.43 (0.86, 2.37) | 0.17 | 1.64 (0.96, 2.81) | 0.07 |
| | Unknown | 1,374 | 72.81 | | | | |
| Sample year | 2009 | 1 | 0.05 | Reference | | Reference | |
| | 2010 | 159 | 8.43 | 1.33 (0.61, 2.92) | 0.48 | 1.48 (0.63, 3.48) | 0.37 |
| | 2011 | 137 | 7.26 | 1.37 (0.62, 3.06) | 0.44 | 1.50 (0.63, 3.58) | 0.37 |
| | 2012 | 146 | 7.74 | 1.4 (0.64, 3.1) | 0.40 | 1.55 (0.66, 3.67) | 0.32 |
| | 2013 | 185 | 9.80 | 1.26 (0.58, 2.74) | 0.55 | 1.43 (0.62, 3.32) | 0.40 |
| | 2014 | 135 | 7.15 | 1.4 (0.63, 3.11) | 0.41 | 1.70 (0.72, 4.04) | 0.23 |
| | 2015 | 188 | 9.96 | 1.5 (0.7, 3.21) | 0.30 | 1.79 (0.79, 4.09) | 0.16 |
| | 2016 | 237 | 12.56 | 1.18 (0.55, 2.51) | 0.67 | 1.27 (0.56, 2.9) | 0.57 |
| | 2017 | 170 | 9.01 | 0.91 (0.4, 2.04) | 0.81 | 1.04 (0.43, 2.5) | 0.94 |
| | 2018 | 215 | 11.39 | 0.89 (0.41, 1.96) | 0.78 | 1.00 (0.42, 2.34) | 1.00 |
| | 2019 | 124 | 6.57 | 0.87 (0.36, 2.06) | 0.74 | 1.02 (0.40, 2.60) | 0.96 |
| | 2020 | 100 | 5.30 | 0.93 (0.38, 2.27) | 0.87 | 1.08 (0.41, 2.83) | 0.87 |
| | 2021 | 12 | 0.64 | 0.62 (0.07, 5.32) | 0.66 | 0.80 (0.09, 6.99) | 0.84 |
| | 2022 | 78 | 4.13 | Empty | | Empty | |
| 65 to 84 | | | | | | | |

| | | | | | | | |
|---|---|---|---|---|---|---|---|
| Gender | Male | 905 | 56.35 | Reference | | Reference | |
| | Female | 696 | 43.34 | 1.06 (0.84, 1.33) | 0.64 | 1.10 (0.86, 1.40) | 0.46 |
| | Unknown | 5 | 0.31 | | | | |
| Ethnicity | White | 1,380 | 85.93 | Reference | | Reference | |
| | Asian | 63 | 3.92 | 0.84 (0.45, 1.56) | 0.58 | 0.89 (0.46, 1.73) | 0.73 |
| | Black | 7 | 0.44 | 0.54 (0.06, 4.47) | 0.57 | 0.70 (0.08, 5.85) | 0.74 |
| | Mixed | 6 | 0.37 | 0.64 (0.07, 5.53) | 0.69 | 0.84 (0.10, 7.23) | 0.88 |
| | Other | 4 | 0.25 | 1.07 (0.11, 10.35) | 0.95 | 1.40 (0.15, 13.54) | 0.77 |
| | Unknown | 146 | 9.09 | | | | |
| IMD | 1 | 179 | 11.15 | Reference | | Reference | |
| | 2 | 202 | 12.58 | 0.64 (0.41, 1.02) | 0.06 | 0.95 (0.57, 1.59) | 0.85 |
| | 3 | 245 | 15.26 | 0.61 (0.39, 0.95) | 0.03 | 0.88 (0.53, 1.44) | 0.60 |
| | 4 | 255 | 15.88 | 0.79 (0.52, 1.21) | 0.28 | 1.27 (0.79, 2.02) | 0.33 |
| | 5 | 242 | 15.07 | 0.70 (0.45, 1.08) | 0.11 | 0.91 (0.56, 1.5) | 0.72 |
| | Unknown | 483 | 30.07 | | | | |
| Lineage | $M1_{global}$ | 184 | 11.46 | Reference | | Reference | |
| | $M1_{13SNP}$ | 10 | 0.62 | 0.67 (0.14, 3.26) | 0.62 | 0.82 (0.17, 4.01) | 0.81 |
| | $M1_{23SNP}$ | 5 | 0.31 | Empty | | Empty | |
| | $M1_{UK}$ | 239 | 14.88 | 1.16 (0.75, 1.77) | 0.51 | 1.17 (0.75, 1.83) | 0.48 |
| | Unknown | 1,168 | 72.73 | | | | |
| Sample year | 2009 | 2 | 0.12 | | | | |
| | 2010 | 131 | 8.16 | Reference | | Reference | |
| | 2011 | 126 | 7.85 | 0.46 (0.03, 7.46) | 0.58 | 0.35 (0.02, 5.76) | 0.46 |
| | 2012 | 120 | 7.47 | 0.37 (0.02, 6.07) | 0.49 | 0.25 (0.01, 4.1) | 0.33 |
| | 2013 | 119 | 7.41 | 0.38 (0.02, 6.24) | 0.50 | 0.32 (0.02, 5.26) | 0.42 |
| | 2014 | 115 | 7.16 | 0.37 (0.02, 6.06) | 0.48 | 0.27 (0.02, 4.4) | 0.36 |
| | 2015 | 148 | 9.22 | 0.26 (0.02, 4.37) | 0.35 | 0.22 (0.01, 3.72) | 0.30 |
| | 2016 | 175 | 10.9 | 0.32 (0.02, 5.27) | 0.43 | 0.23 (0.01, 3.85) | 0.31 |
| | 2017 | 162 | 10.09 | 0.37 (0.02, 5.99) | 0.48 | 0.32 (0.02, 5.16) | 0.42 |
| | 2018 | 232 | 14.45 | 0.30 (0.02, 4.85) | 0.39 | 0.18 (0.01, 3.01) | 0.23 |
| | 2019 | 128 | 7.97 | 0.22 (0.01, 3.61) | 0.29 | 0.19 (0.01, 3.1) | 0.24 |
| | 2020 | 85 | 5.29 | 0.21 (0.01, 3.45) | 0.27 | 0.14 (0.01, 2.4) | 0.18 |
| | 2021 | 3 | 0.19 | 0.49 (0.03, 8.15) | 0.62 | 0.42 (0.03, 6.93) | 0.54 |
| | 2022 | 60 | 3.74 | 0.50 (0.01, 19.56) | 0.71 | 0.50 (0.01, 19.56) | 0.71 |
| 85 and above | | | | | | | |
| Gender | Male | 210 | 41.58 | Reference | | Reference | |
| | Female | 294 | 58.22 | 1.44 (1.01, 2.06) | 0.04 | 1.21 (0.84, 1.74) | 0.30 |

| | | | | | | | |
|---|---|---|---|---|---|---|---|
| | Unknown | 1 | 0.2 | | | | |
| Ethnicity | White | 444 | 87.92 | Reference | | Reference | |
| | Asian | 10 | 1.98 | 0.69 (0.19, 2.48) | 0.57 | 1.00 (0.28, 3.58) | 1.00 |
| | Black | 2 | 0.40 | 1.04 (0.06, 16.68) | 0.98 | 1.49 (0.09, 24.05) | 0.78 |
| | Mixed | 0 | | Empty | | Empty | |
| | Other | 0 | | Empty | | Empty | |
| | Unknown | 49 | 9.70 | | | | |
| IMD | 1 | 65 | 12.87 | Reference | | Reference | |
| | 2 | 62 | 12.28 | 0.91 (0.45, 1.82) | 0.79 | 0.69 (0.34, 1.4) | 0.30 |
| | 3 | 73 | 14.46 | 1.18 (0.6, 2.3) | 0.64 | 1.07 (0.55, 2.1) | 0.83 |
| | 4 | 76 | 15.05 | 0.78 (0.4, 1.52) | 0.48 | 0.80 (0.41, 1.57) | 0.52 |
| | 5 | 85 | 16.83 | 1.61 (0.83, 3.09) | 0.16 | 0.99 (0.52, 1.89) | 0.97 |
| | Unknown | 144 | 28.51 | | | | |
| Lineage | $M1_{global}$ | 46 | 9.11 | Reference | | Reference | |
| | $M1_{13SNP}$ | 2 | 0.40 | 0.84 (0.05, 14.26) | 0.90 | 1.09 (0.06, 18.51) | 0.95 |
| | $M1_{23SNP}$ | 4 | 0.79 | 0.84 (0.11, 6.49) | 0.87 | 0.36 (0.04, 3.76) | 0.40 |
| | $M1_{UK}$ | 93 | 18.42 | 0.86 (0.42, 1.74) | 0.67 | 0.66 (0.32, 1.35) | 0.25 |
| | Unknown | 360 | 71.29 | | | | |
| Sample year | 2009 | 0 | | | | | |
| | 2010 | 29 | 5.74 | Reference | | Reference | |
| | 2011 | 29 | 5.74 | 0.57 (0.2, 1.62) | 0.29 | 0.76 (0.27, 2.13) | 0.60 |
| | 2012 | 41 | 8.12 | 0.78 (0.3, 2.06) | 0.62 | 0.85 (0.33, 2.22) | 0.74 |
| | 2013 | 40 | 7.92 | 0.55 (0.21, 1.46) | 0.23 | 0.44 (0.16, 1.16) | 0.10 |
| | 2014 | 43 | 8.51 | 0.58 (0.22, 1.52) | 0.27 | 0.59 (0.23, 1.51) | 0.27 |
| | 2015 | 61 | 12.08 | 0.59 (0.24, 1.46) | 0.25 | 0.56 (0.23, 1.38) | 0.21 |
| | 2016 | 62 | 12.28 | 0.39 (0.16, 0.96) | 0.04 | 0.31 (0.12, 0.77) | 0.01 |
| | 2017 | 47 | 9.31 | 0.69 (0.27, 1.78) | 0.45 | 0.66 (0.26, 1.66) | 0.38 |
| | 2018 | 48 | 9.5 | 0.86 (0.33, 2.2) | 0.75 | 0.75 (0.3, 1.89) | 0.54 |
| | 2019 | 43 | 8.51 | 0.33 (0.12, 0.87) | 0.03 | 0.31 (0.12, 0.85) | 0.02 |
| | 2020 | 41 | 8.12 | 0.86 (0.33, 2.28) | 0.77 | 0.77 (0.3, 2.01) | 0.60 |
| | 2021 | 6 | 1.19 | 1.22 (0.19, 7.82) | 0.83 | 1.63 (0.26, 10.32) | 0.61 |
| | 2022 | 15 | 2.97 | 0.31 (0.08, 1.13) | 0.08 | 0.20 (0.05, 0.88) | 0.03 |

### Supplementary Figure S1

**A.**

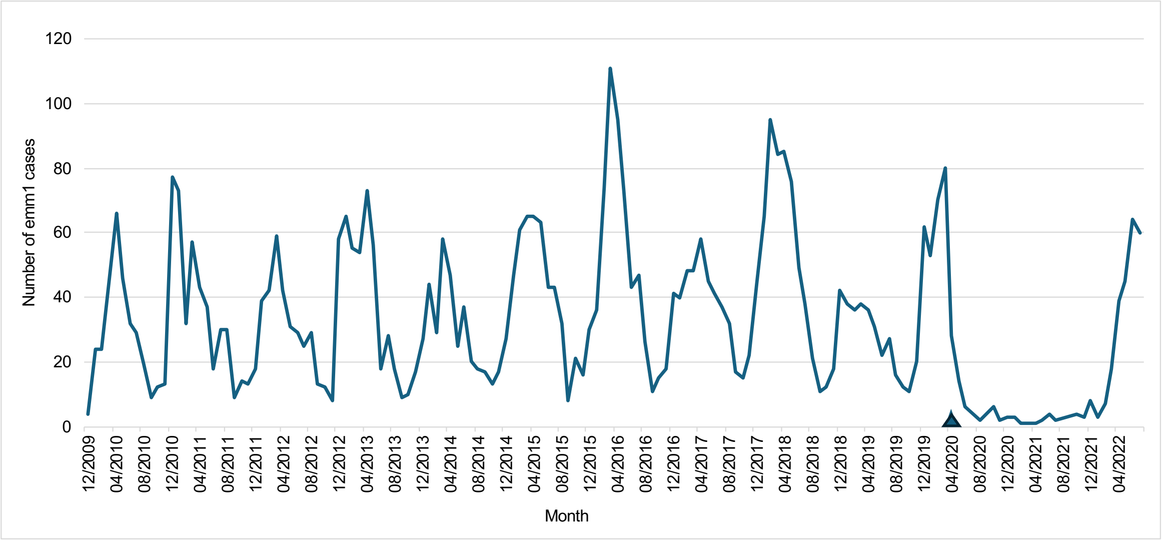


**B.**

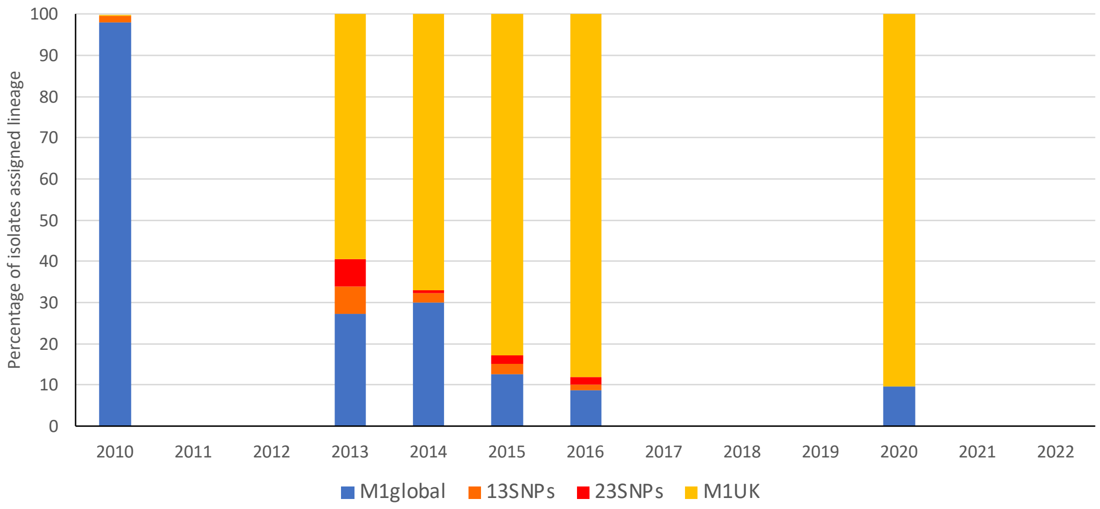


**Supplementary Figure S1 Proportion of emm1 iGAS isolates assigned to lineage per year.** Total iGAS cases associated with *emm*1 *S. pyogenes* isolates per month referred to the reference laboratory are shown over the study period 2010-2022. UK lockdown onset indicated with triangle (A). Proportion of isolates per year that were used for lineage assignment, with lineage indicated (B).

### Supplementary Figure S2

A.

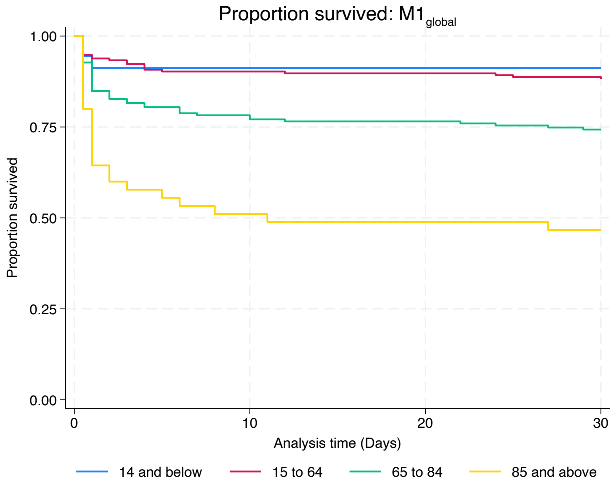


B.

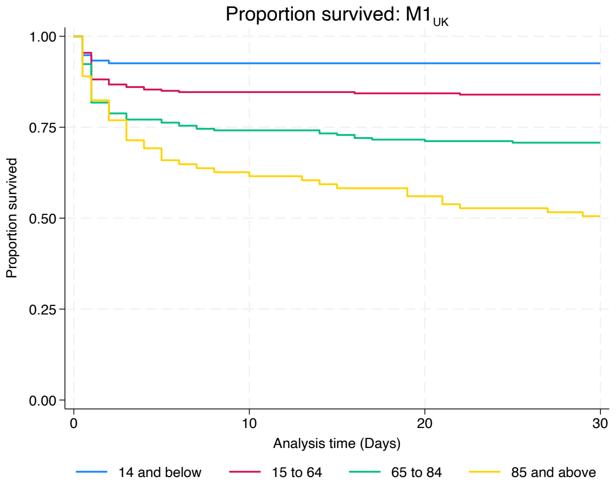


**Supplementary Figure S2. Kaplan Meier survival plot showing 30-day survival of $M1_{global}$ and $M1_{UK}$ iGAS cases by age group.**

Left panel (A) shows $M1_{global}$ iGAS cases. Right panel (B) shows $M1_{UK}$ iGAS cases. Age group indicated by line colour. Children aged under 15y blue; Adults aged 15y - 64y red; Adults aged 65y -8y4 green; Adults aged over 85y yellow.

### Supplementary Figure S3

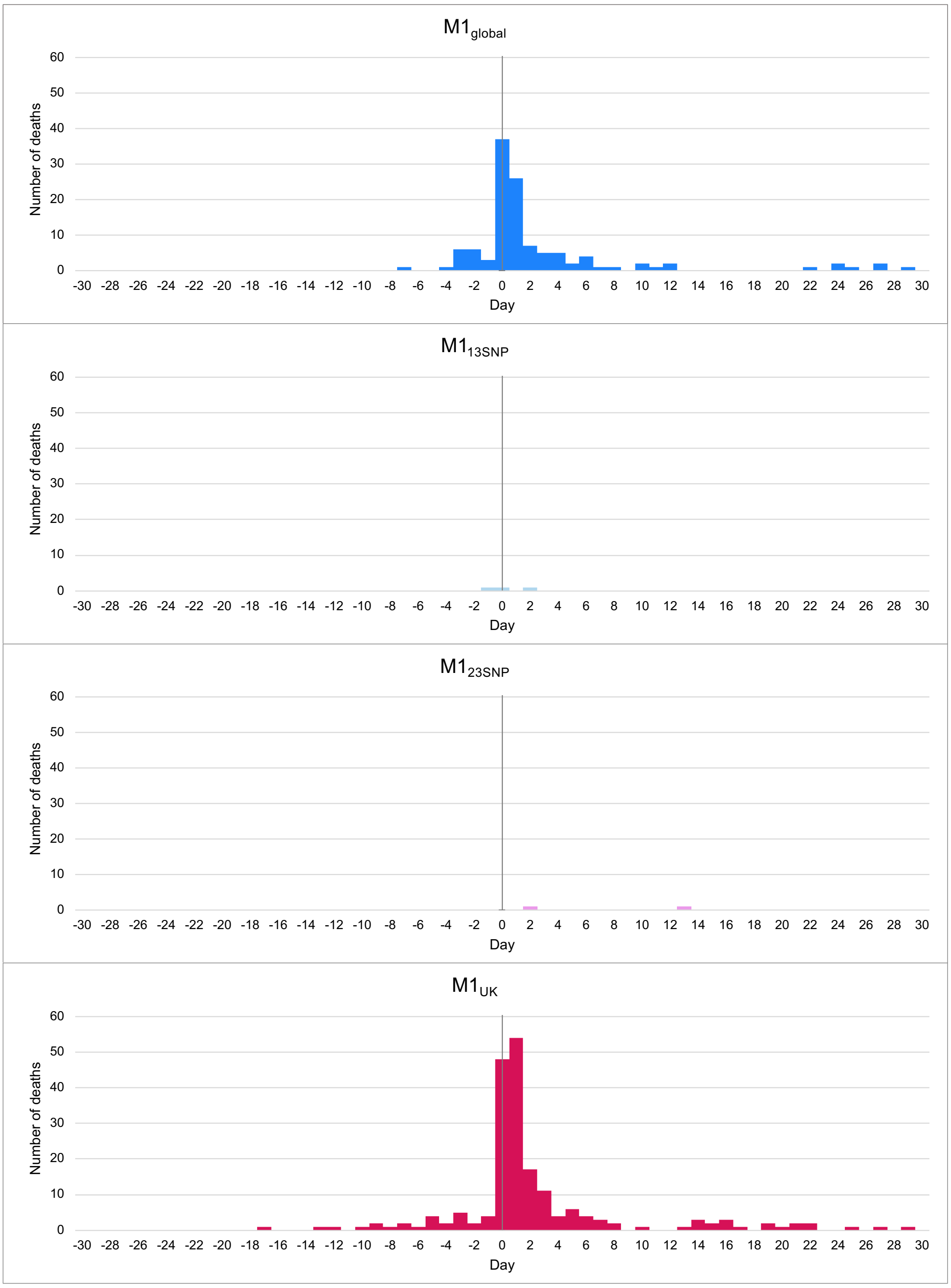


**Supplementary Figure S3. Time between diagnostic sample and death by lineage.** Each panel shows deaths from each *emm*1 lineage as labelled from top to bottom: $M1_{global}$ (blue); $M1_{13SNPs}$ (light blue); $M1_{23SNPs}$ (light pink); $M1_{UK}$ (red). Y axes show numbers of deaths for each sublineage. X axes show number of days since date of diagnostic sample that yielded *S. pyogenes*.
